# Comparing Five Generative AI Chatbots’ Answers to LLM-Generated Clinical Questions with Medical Information Scientists’ Evidence Summaries

**DOI:** 10.1101/2025.09.24.25336199

**Authors:** Mallory N. Blasingame, Taneya Y. Koonce, Annette M. Williams, Jing Su, Dario A. Giuse, Poppy A. Krump, Nunzia B. Giuse

## Abstract

**Objective:** To compare answers to clinical questions between five publicly available large language model (LLM) chatbots and information scientists.

**Methods:** LLMs were prompted to provide 45 PICO (patient, intervention, comparison, outcome) questions addressing treatment, prognosis, and etiology. Each question was answered by a medical information scientist and submitted to five LLM tools: ChatGPT, Gemini, Copilot, DeepSeek, and Grok-3. Key elements from the answers provided were used by pairs of information scientists to label each LLM answer as in Total Alignment, Partial Alignment, or No Alignment with the information scientist. The Partial Alignment answers were also analyzed for the inclusion of additional information.

**Results:** The entire LLM set of answers, 225 in total, were assessed as being in Total Alignment 20.9% of the time (n=47), in Partial Alignment 78.7% of the time (n=177), and in No Alignment 0.4% of the time (n=1). Kruskal-Wallis testing found no significant performance difference in alignment ratings between the five chatbots (*p*=0.46). An analysis of the partially aligned answers found a significant difference in the number of additional elements provided by the information scientists versus the chatbots per Wilcoxon-Rank Sum testing (*p*=0.02).

**Discussion:** Five chatbots did not differ significantly in their alignment with information scientists’ evidence summaries. The analysis of partially aligned answers found both chatbots and information scientists included additional information, with information scientists doing so significantly more often. An important next step will be to assess the additional information both from the chatbots and the information scientists for validity and relevance.

## Introduction

Generative artificial intelligence (AI) tools are increasingly embedded into the systems and workflows used by experts and the general public to search for health information. In 2024, a Kaiser Family Foundation poll of 2,428 U.S. adults found that roughly 1 in 6 (17%) respondents used AI chatbots at least monthly to seek out health information [1]. Even when searchers do not directly query a generative AI tool, they may increasingly encounter large language model (LLM)-generated answers as they search for health information on the web [2] or in proprietary literature databases [3,4]. Google and Microsoft Bing are now providing generative AI answers in search results, including responses to medical queries [5]. These features provide users with quick, easy access to synthesized information, which may be useful for guiding conversations with clinical providers [6] but may also pose unforeseen risks [7,8].

National Center for Health Statistics data from 2022 revealed that 58.5% of adults surveyed had looked for health information on the Internet in the past year [9], and Google reported in 2019 receiving more than 1 billion health-related searches per day [10]. Thus, it is likely that many people are encountering generative AI answers to their everyday health inquiries. It is important to understand how these answers compare to information provided by highly trained, trusted information scientist professionals with experience and formal training in medical librarianship. Previous studies have investigated LLMs’ performance for answering medical questions using a variety of study designs and evaluation dimensions [11] and assessed their ability to aid with steps of the evidence synthesis process including search strategy development [12–14] and systematic review tasks such as citation screening and data extraction [15,16]. However, additional investigation is needed into how generative AI chatbots’ answers to medical questions compare with actual evidence synthesis responses developed by information scientists. To address this knowledge gap, our team at the Vanderbilt University Medical Center (VUMC) Center for Knowledge Management (CKM) has embarked on a series of studies to investigate generative AI in the context of medical information sciences. In a previous study, we assessed the performance of VUMC’s internally managed version of GPT-4 (aiChat) [17], using medical information scientists’ evidence summaries as the gold standard for comparison [18]. In that initial study, we compared aiChat answers with summaries our team previously developed in response to a VUMC-proprietary set of questions received during rounds or from an electronic health record-linked message basket. The study revealed that 83.3% of aiChat responses included all of the elements from the information scientist summaries that were identified as being most critical for answering the questions, while also reflecting known limitations of generative AI tools, including fabrication of references. In a second study comparing search strategies generated by three publicly available LLMs with information scientists’ expert searches, our team found the AI chatbots were able to generate Boolean search queries but missed many relevant keywords and often included inaccurate controlled vocabulary terms [19].

As a next phase in our generative AI research agenda, the current study aimed to build on these findings by conducting a prospective, in-depth, detailed comparison between five generative AI chatbots’ answers to clinical questions and medical information scientists’ evidence syntheses, focusing on tools that are widely available and likely to be used by the public. To avoid using proprietary questions in these publicly available systems, we prompted the chatbots to create the questions for the study, and information scientists developed new evidence syntheses for each question to compare with the LLMs’ answers. As the summaries created by the information scientists were not reviewed and validated by clinical experts, we did not consider them to be a “reference standard” for assessing accuracy but rather compared them one-to-one with the LLM answers. The study specifically investigated the following questions:

1. How do the answers provided by five publicly available LLMs compare to those of medical information scientists?
2. Are there differences in the alignment with information scientist answers when the AI tools answer their own generated questions?

## Methods

The study received a non-human subjects research determination from the VUMC Institutional Review Board (IRB #241743). The reporting of this study follows the Chatbot Assessment Reporting Tool (CHART) guidelines [20,21]; the CHART Methodological Diagram and Checklist can be viewed in Appendix A.

### Generative AI Chatbots

When the study was initially conceived in October 2024, ChatGPT (https://chatgpt.com/), Google Gemini (https://gemini.google.com/), and Microsoft Copilot (https://copilot.microsoft.com/) were selected for investigation based on their reported frequency of use in medical literature to date [22], and, in the cases of Gemini and Copilot, their increasing integration into highly used public search engines. In early 2025, as study activities were ongoing, the release of DeepSeek (https://chat.deepseek.com) and Grok-3 (https://grok.com/) for public use sparked a great deal of interest; thus, we decided to add them to the analysis to explore any differences in performance with these newer models. The free, public, web-based versions were used; in Google Gemini, DeepSeek, and Grok-3, logging into a free personal account was required. The versions used were the most current freely-available, base models for each tool at the time: ChatGPT-4o [23], Gemini 2.0 Flash [24], DeepSeek R1 [25], and Grok-3 [26] with web search enabled; the particular model of Copilot was not specified in the system. DeepSeek defines itself as an open-source LLM; all other models used in this study are closed-source.

### Prompt Engineering

Three prompts were used in the study: 1) a prompt to obtain the questions from the generative AI chatbots; 2) a prompt to obtain the *answers* from the LLMs; and 3) a prompt to submit each information scientist and chatbot answer to the generative AI tools to obtain lists of key elements for comparison. The prompt for obtaining the LLM answers was reused from our previous study [18]. The other two prompts were newly created for this study by the co-authors, who include individuals with formal training and expertise in information sciences, medicine, public health, and biomedical informatics. The COSTAR framework [27] was followed, with each prompt including a section on Context, Objective, Style, Tone, Audience, and Response. When possible, elements of the original prompt were reused, with adjustments made to tailor the prompt to the specific task. The prompts were submitted to the tools for testing and revised as needed. In all cases, a new session was started with the chatbot for each individual prompt submitted.

### Obtaining the Questions

In our previous study [18], we used questions received from clinicians based on actual clinical encounters; these questions are proprietary to our institution and thus could only be used with our organization’s internally managed AI tool. For this study that uses publicly available LLMs, we intentionally used the chatbots to create non-proprietary clinical questions that could be input into the tools and publicly shared (see Appendix B). In evidence-based medicine, the use of well-structured questions can enhance precision and aid in establishing discrete concepts for search strategy formulation and information retrieval [28]. Since the 1990s, the PICO (patient, intervention, comparison, outcome) framework has been used by clinicians and information scientists to guide evidence searching, filtering, and selection in response to both clinical questions and inquiries from members of the lay public [29]. Thus, we leveraged the LLMs’ ability to quickly and efficiently generate PICO-structured questions for use in this study; each question was subsequently reviewed by an information scientist with formal education and training in medicine to ensure all questions generated were medically plausible.

Forty-five questions were obtained in November 2024 by prompting ChatGPT, Google Gemini, and Microsoft Copilot to each provide five treatment questions, five prognosis questions, and five etiology questions in PICO format (Appendix B). DeepSeek and Grok-3 were not used to generate questions, as the tools were not yet widely available for public use. The question categories were based on the most common types of questions our team has received in our history of providing clinical evidence services [18]. After initial prompt testing revealed that the tools tended to provide questions focused on treatment even when asked for prognosis or etiology questions, definitions of prognosis and etiology were added to the prompts for these categories (Appendix C).

The questions were obtained by two information scientists, one of whom has medical training and reviewed each set of generated questions for fit with the designated category and clinical plausibility. When duplicate or non-medically plausible questions were generated, they were removed from the question pool and the tool that provided the question was prompted for an additional question as part of the same session. For example, one generated question asked about a patient population with Type 2 diabetes whose HbA1c level was within a “normal range at diagnosis”; given elevated HbA1c is a standard diagnostic criterion for Type 2 diabetes [30], we excluded this question and prompted the chatbot to provide another.

### Question Assignment

Questions were assigned to four highly trained information scientists with experience developing evidence syntheses in response to clinical queries. The number of questions given to each information scientist was based on the amount of effort each had assigned to the project. The questions were initially randomly assigned with stratification by the chatbot from which they originated and question category. However, as efforts shifted throughout the project period, five questions were reassigned. Ultimately, two information scientists answered 13 questions each, one information scientist answered 14 questions, and one information scientist answered five questions.

### Developing the Information Scientist Evidence Summaries

Our team’s standard practices for developing evidence syntheses were followed. As often done at our center, information scientists individually developed comprehensive PubMed search strategies for each question and then met as a group to review the searches and provide feedback on areas for refinement. Once the searches were finalized, the information scientists completed evidence summaries for their questions, following the template used by our team for answering real-world evidence queries. The template reflected changes adopted by the team after our previous AI study [18] and includes the following sections: 1) an introduction, including a brief summation of findings, characterization of the state of the literature, and definition of key topics; 2) a summary of selected literature, including the design, publication year, aims, and results for each selected study; and 3) conclusion, with the “bottom line” of findings from the literature and a brief summary of strengths and limitations. After each summary was completed, it was stored in REDCap [31,32]. A second information scientist reviewed each summary to confirm alignment with the template and that all required fields (such as the date the packet was finalized and summary supporting references) were completed.

### Obtaining Answers from the Generative AI Tools

As the information scientists finalized each summary, a senior information scientist not involved in evidence synthesis submitted the corresponding question to each chatbot using a standardized prompt (Appendix D). In brief, the prompt asked the tools to provide an evidence summary in response to the provided clinical question in “the role of a medical librarian,” with the answer limited to the information available as of the date the information scientist completed the summary. All information scientist summaries were completed and answers captured from the generative AI tools between January and April 2025. Each response was stored in full in the study database, along with references and any hyperlinks included with the response.

### Comparing the Information Scientist Evidence Summaries with Generative AI Answers

In our research assessments of generative AI tools, CKM recognized both the value of the LLMs to clearly and effectively summarize evidence and, when requested, organize the information into easy-to-understand key elements. Leveraging on this understanding and with the intent to remove human subjectivity, for the current study, the team decided to have each of the five LLMs automatically generate (Appendix E) the key elements for the LLM-generated 45 summaries and the 45 summaries generated by the information scientists. Each set of key elements from all the summaries, whether generated by the LLMs or written by the information scientists, was subsequently reviewed by information scientists to ensure the key elements accurately reflected the summary.

Because of the lack of direct involvement and interaction with clinicians, a decision was also made not to label the information scientists’ summaries as “reference standards.” Without the opportunity for consultation, we cannot assume that information scientists and clinical experts will agree on the answers to clinical questions. A study by Tao and colleagues found that librarians and healthcare professionals differed significantly in their ratings of the comprehensiveness of ChatGPT responses [33].

Given the above consideration, agreement and disagreement among the key elements was used to determine the level of alignment between the answers of the LLMs and information scientists; the analysis was conducted by four unblinded information scientists. A pair of information scientists reviewed and gathered consensus on whether the information included in the key elements from the answers of each of the five LLM tools – ChatGPT, Gemini, Copilot, DeepSeek, and Grok-3 – was a) totally aligned, b) partially aligned, or c) not aligned with the information expressed by the key elements from the information scientists’ answers. In each of the above instances, one of the information scientists in the pair was the author of the summary compared.

We used the concept of *Total Alignment* when the information expressed by the key elements in each of the answers being compared was judged as being the same. Although the lists of key elements were numbered, the summaries did not need to have the same number of elements to be considered in Total Alignment; the elements simply provided a quick and straightforward means for assessing whether the content in the answers agreed.

For example, a single key element from the information scientist’s response could be determined to align with two or more elements from the chatbot’s response, and vice versa. Additionally, differences in wording or data cited that represented the same concept or viewpoint were counted as in alignment. *Partial Alignment* included four sub-categories:

1) All key elements expressing the same concepts were in agreement, but the *tool’s* answer included additional key elements; 2) All key elements expressing the same concepts were in agreement, but the *information scientist’s* answer included additional key elements; 3) All key elements expressing the same concepts were in agreement, but *both* answers included additional key elements; or 4) The information scientist and LLM agreed on some but not all key elements expressing the same concepts, and one or both answers may have included additional key elements. For categories 1-3, we tabulated how many additional key elements were provided by the LLM or the information scientist. *No Alignment* indicated that none of the key elements agreed.

### Sample Size Determination

The sample size was informed by the need to obtain multiple questions from each of the three tools we originally intended to study (ChatGPT, Gemini, and Copilot) and the three categories selected for the questions (treatment, prognosis, and etiology). Consideration was also given to the estimated time (historically averaging about 8 hours per summary [34]) needed by the information scientists to generate summaries, a consideration largely dictated by their availability. All of this brought us to the final determination that using a minimum of five questions for each of the question categories (total of 15 questions) for each of the three tools could give us adequate data for our study. A formal sample size calculation was not conducted.

### Data Analysis

Descriptive statistics were used to report the frequency and proportion of responses assessed as in Total Alignment, Partial Alignment, or No Alignment for each of the tools in comparison with the information scientist responses, and to characterize the number and percentage of each type of Partial Alignment by tool. A Kruskal-Wallis test was used to assess whether there was a significant difference in the five generative AI tools’ alignment with the information scientists’ responses, and in the types of partial alignment across the five tools. This test was selected as the comparison focused on multiple independent groups and the data were nonparametric. For the partially aligned answers in categories 1-3 above, the median number and mean proportion of key elements identified as “additional” from the information scientist and LLM were calculated. A Wilcoxon Rank-Sum test was used to compare the two groups of data on the number of additional key elements from the information scientist summaries versus the chatbot summaries. In a sub-analysis focused on the three generative AI tools that provided the PICO questions (ChatGPT, Gemini, and Copilot), a Friedman statistic test was used to assess whether there were any significant differences in each of the three tools’ alignment based on whether they were answering their own questions or those provided by the other two chatbots. The Friedman statistic test was chosen due to the use of dependent groups and rank-based data. All statistical analyses were conducted in GraphPad Prism, and visualizations were created in Flourish and Microsoft Excel.

## Results

In total, 53 PICO questions were generated by ChatGPT, Gemini, and Copilot. During the process of generating each initial set of 15 questions from the three tools, eight questions were removed from the pool after review by an information scientist with medical expertise and eight new questions generated, resulting in the final set of 45 questions. Reasons for question exclusion included determination that the question was clinically implausible (n=3; 2 from ChatGPT, 1 from Gemini), that the question was a duplicate of another question already in the pool (n=4; 2 from Copilot, 2 from ChatGPT), or that the question did not align with the requested category (n=1 from ChatGPT).

### Alignment between the Information Scientist and Generative AI Responses

Across the 225 answers generated to the 45 questions by the five generative AI tools, 47 (20.9%) were in Total Alignment with the information scientist’s response, 177 (78.7%) were in Partial Alignment, and one (0.4%) was assessed as having No Alignment. The response with No Alignment was from ChatGPT. Gemini had the highest frequency of Total Alignment ratings (13/45; 28.9%), while Grok-3 had the lowest frequency of answers in Total Alignment (7/45; 15.6%). A Kruskal-Wallis test revealed no significant differences in the alignment ratings between the five tools (*p*=0.46). The full alignment ratings for each chatbot can be viewed in Figure 1.

**Figure 1.**
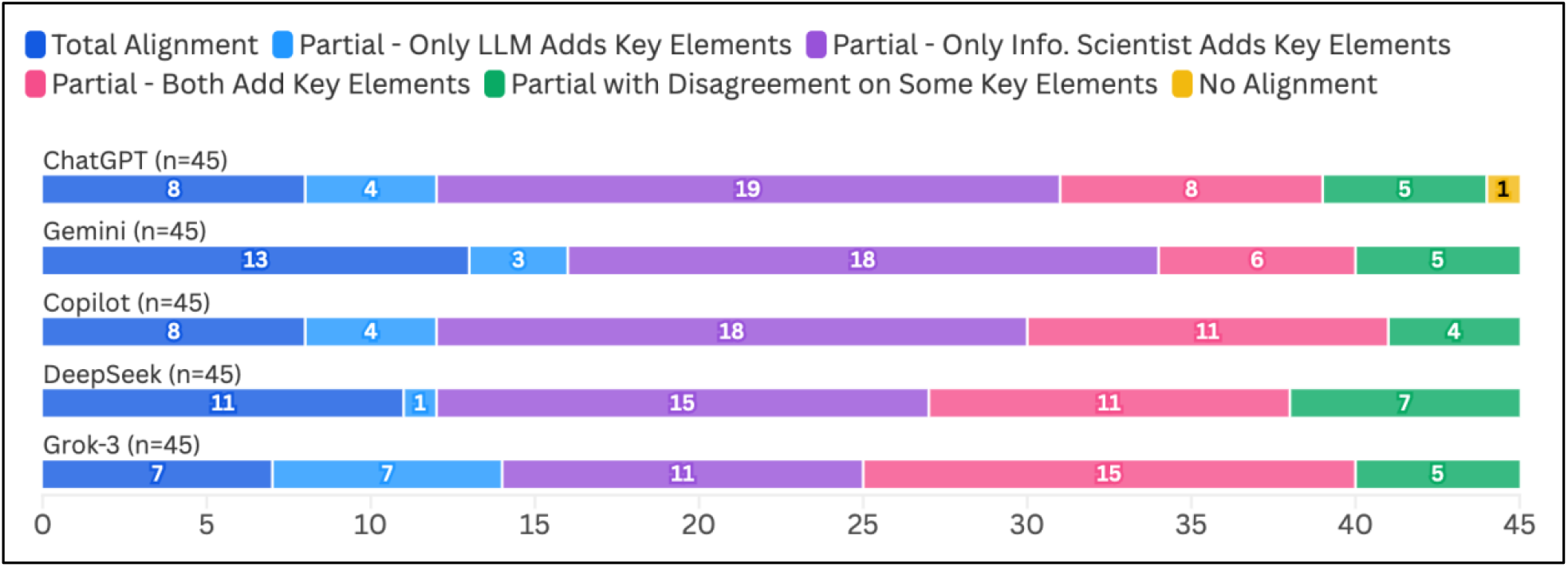
Alignment between Information Scientist and LLM Answers (N=225)

### Analysis of Partially Aligned Responses

In the subset of 177 responses that we labeled as in Partial Alignment, 151 (85.3%) had all key elements expressing the same concepts in agreement, but the answers from the generative AI tool (n=19), the information scientist (n=81), or both (n=51) contained additional unique elements. The remaining 26 (14.7%) answers labeled as Partial Alignment agreed only on some of the key elements expressing the same concepts; in all cases in this category, there was only one concept identified as being in disagreement. Grok-3 had the highest frequency of responses in which only the LLM included additional key elements (7/45; 15.6%) or both the LLM and the information scientist provided additional key elements (15/45; 33.3%). ChatGPT had the most answers for which only the information scientist included additional key elements (19/45; 42%), and DeepSeek had the most answers that did not agree on all key elements in common with the information scientist’s answer (7/45; 15.6%). However, a Kruskal-Wallis analysis found no significant differences in type of Partial Alignment among the five tools (*p*=0.78). Full Partial Alignment results by tool can be viewed in Figure 1.

### Analysis of Additional Key Elements

The median number of total elements from the *chatbot* answers providing additional information (total of 70) was 5.5 (range of 3-8). There was a median of 1 (range: 1-3) additional key element per answer. The median number of total elements from the *information scientists’* answers with additional key elements (total of 132) was 5 (range: 3-10). There was a median of 1 (range: 1-4) additional key element per answer. The distribution of the number of additional key elements identified in each information scientist and chatbot answer in this category is shown in Figure 2. A Wilcoxon Rank-Sum test found a significant difference when comparing the groups of additional key elements generated for the chatbot summaries versus the information scientist summaries (*p*=0.02).

**Figure 2.**
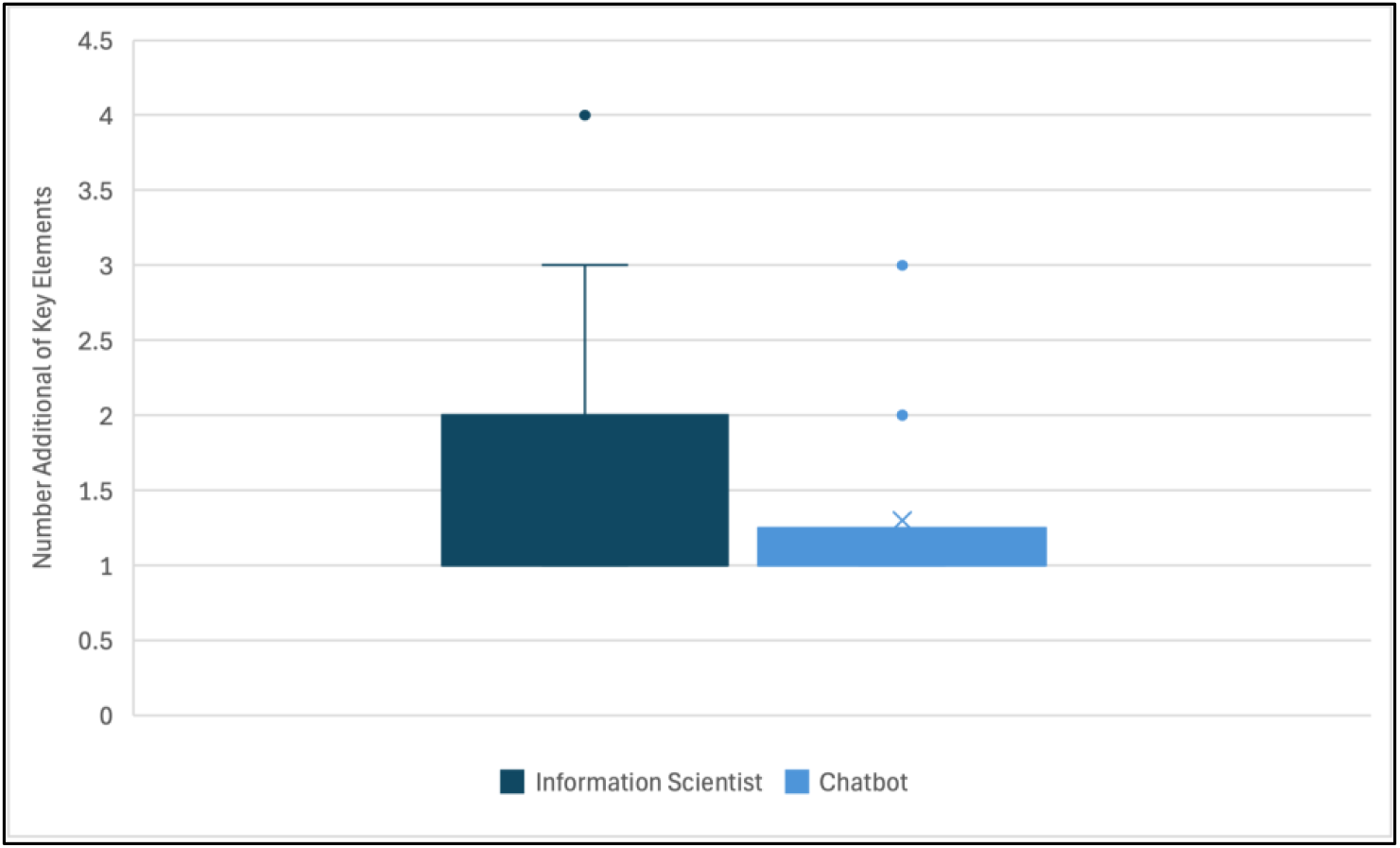
Distribution of Additional Key Elements from the Information Scientist (n=132) and Chatbot Answers (n=70)

### Alignment by Question Source

In the sub-analysis limited to the three LLMs that provided the questions (ChatGPT, Gemini, and Copilot), all three of the chatbots had the highest frequency of Total Alignment ratings when answering Copilot questions (Figure 3). However, per Friedman statistic testing, no significant differences in alignment ratings were found for any of the three tools when comparing the alignment ratings of their answers to their own questions against the alignment ratings of their answers to the other two tools’ questions (*p*=0.24 for ChatGPT, 0.88 for Gemini, and 0.56 for Copilot). Full alignment ratings by question source for these three tools can be viewed in Figure 3.

**Figure 3.**
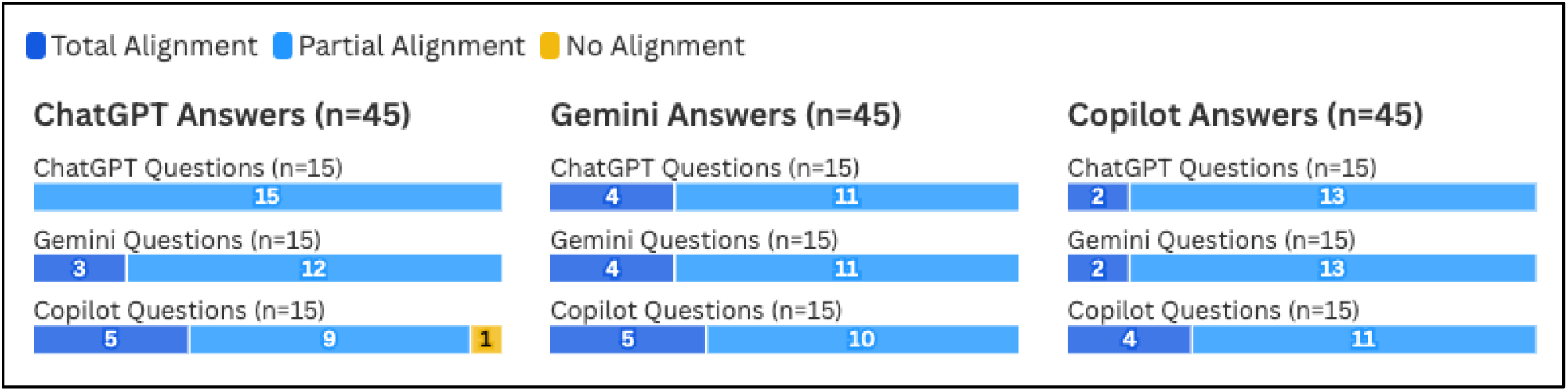
Alignment between Information Scientist and LLM by Individual Chatbot (ChatGPT, Gemini, and Copilot)

### Descriptive Characteristics of the Information Scientist and Generative AI Summaries

On average, the information scientists’ summaries were longer than the generative AI answers, with a mean of 1,668 words observed for the information scientists’ answers compared with average word counts ranging from 286 (Copilot) to 497 words (Grok-3) for the generative AI tools’ answers. The generative AI tools commonly included direct links to references cited in the summaries (100% of ChatGPT answers had linked references, 91% of Gemini answers, 84% of Copilot answers, and 60% of Grok-3 answers), except for DeepSeek, which did not provide direct links to cited references. Consistent with our team’s standard practice, all 45 information scientist summaries included full references with hyperlinks.

## Discussion

This study is the second in what we envision being a step-by-step series to clearly investigate all elements of AI performance as applicable to the functional work of information scientists. As we wanted to create an optimal scenario for question answering, we intentionally had the LLMs generate PICO-formatted questions that were then verified by an information scientist with medical training. Other authors such as Demir and colleagues (2024) have examined LLMs’ ability to generate PICO-formatted questions [35]. While the PICO format provides a useful and well-established framework to prompt LLMs to generate questions in alignment with prompt engineering techniques [36], it is not always characteristic of questions generated in fast-track clinical settings. As illustrated by a study from Huang and colleagues (2006), clinicians’ questions are often missing one or more of the PICO elements; of 59 questions posed by clinicians, only two included all four PICO elements, and 37.3% included just the intervention and outcome elements [37]. The number of PICO elements included in the questions impacts the search strategy and scope of the literature and may translate into a different type of literature and resulting summary. Nevertheless, the PICO format is one of the most common mechanisms librarians use to fully research and investigate questions they receive from their users, making this type of exploration highly relevant for understanding the potential role of AI usage in medical librarianship.

Our comparison between information scientists’ evidence syntheses and five generative AI chatbots revealed that in most instances (20.9% total and 78.7% partial), there is a high level of similarity among the summaries reported, even though an in-depth analysis of the partial answers shows that information scientists are more likely to include additional information (key elements) in their answers. Key to this study is the reporting of additional key elements observed in either the large language models’ or the information scientists’ summary responses and the analysis comparing information scientists’ answers to five distinct LLMs. No significant differences were found in the degree to which the five generative AI tools’ responses aligned with the information scientists’ answers, and no impact on performance was observed when the tools answered their self-generated questions, suggesting that the study results were not influenced by whether the questions were based on the LLM’s own training data.

It is worth noting that other studies have found significant differences in LLMs’ answers to clinical questions, although variation in LLMs, question types, and overall methodology makes it difficult to directly compare the existing literature [11]. For example, Lin and colleagues (2025) compared several ChatGPT, Gemini, and Copilot models’ answers to questions on postmenopausal osteoporosis with guideline recommendations and found that ChatGPT-4o’s answers were significantly more accurate than other models [38]. Flaharty et al. (2024) investigated several open- and closed-source models’ performance (including Gemini and ChatGPT-3.5 and 4) in answering genetic questions and found that ChatGPT-4 had the highest performance in terms of correctly identifying a genetic condition, with significant differences observed in several models’ performance depending on whether the question was asked in medical or lay language [39].

We conducted an in-depth analysis of the only “non-alignment” answer in the study (generated by ChatGPT). The answer was reporting on a question asking about the use of early ischemic changes for prediction of “functional outcomes” in acute ischemic stroke patients. In comparing the two responses, we noted that the ChatGPT answer included a reference supporting the opposite viewpoint from the information scientist’s answer. The answer, although not aligned with the information scientist, was reported as one of the multiple viewpoints provided by the other LLMs, giving us a moment of pause and reinforcing our belief that all additional elements provided by LLMs deserve further investigation.

Our study also found the information scientists’ answers more commonly provided additional information than the generative AI chatbots’ responses. This difference may be explained in part by the fact that information scientists’ evidence summaries were longer on average than the LLMs’. It is however notable that, despite their shorter length, 70/225 (31.1%) generative AI summaries were found to provide key elements not present in the information scientists’ summaries. This finding presents a potential for chatbots to serve a complementary role in evidence synthesis by surfacing additional supporting information and/or alternate viewpoints for the information scientist to investigate, verify, and consider for inclusion.

As noted previously, we intentionally did not make a judgment on which answer was correct but rather characterized the answers in terms of alignment between the chatbot and human, with no true “reference standard.” Evidence synthesis is a complex task often involving interaction through the reference interview and ongoing consultation between the subject matter expert and information professional. Hripcsak and Wilcox (2002), in their discussion of reference standards for evaluations of informatics systems, state, “For more complex reasoning tasks, experts are needed to judge the appropriateness of system responses,” and an answer may be assessed by a clinical expert as “appropriate even if it matches none of the comparison responses (e.g., a reasonable medication alternative)” [40]. When a true reference standard is lacking, answers prepared by a human can be compared with answers from the system for “similarity” instead of “performance” [40], consistent with the approach used in our study.

The need to carefully review LLM responses for accuracy is well-established and critical to their use [41]. As evidenced by the above example, chatbots may also introduce additional information from legitimate sources that cannot be assumed to be incorrect without further examination. Humans may also miss relevant information or have differing assessments of relevance depending on their role or area of specialization [35,36]. The focus on alignment and additional information provided by both human and chatbot in this study highlights the potential for LLMs to serve as an aid in a model where the information scientist prepares an answer to an evidence inquiry but uses the chatbot to identify additional information for further investigation and inclusion in her citations and summaries.

These findings build on our approach, throughout our series of studies, of applying a “growth mindset” to the investigation of how AI can be applied to our workflows as information scientists [42]. Similar to the process of consulting a colleague and deciding if and how to incorporate their ideas and feedback, consulting a chatbot can help stimulate our thinking and uncover previous knowledge gaps. By identifying LLMs’ areas of strength and applying them to our work as information scientists, we can begin to truly partner with AI to enhance our performance beyond only productivity gains [42].

### Limitations

Limitations of the study include not assessing the chatbot answers for potential harm or misleading statements. The next planned phase – examine the additional information from the chatbots – will be an important step for further elucidating any potential unfounded information that may have been included in their responses.

Additionally, the information scientist who answered each question was included in the pair of reviewers who assessed the chatbots’ answers, and the reviewers were not blinded to the source of the answers. While this was done intentionally to leverage the information scientist’s knowledge of the topic, it is possible this approach introduced bias.

Four of the chatbots included in the study are closed-source models; thus, reproducibility is limited by the lack of transparency into the models’ training sets and design. We did not conduct a formal investigation into reproducibility of chatbot answers. We also acknowledge that as time passes and models are updated, reproducibility may be limited as the chatbots have access to more data, due to both the emergence of new knowledge and additions to the LLMs’ training sets. Stanford’s Holistic Evaluation of Language Models for Medical Tasks (medHELM) Leaderboard demonstrates that performance of LLMs on medical domain scenarios varies between model versions, with Gemini, for example, showing improved accuracy with 2.0 Flash versus 1.5 Pro [43].

Finally, the comparison of the LLM and information scientist answers relied on the key elements selected by each tool. The information scientists agreed that the elements selected were appropriate, but we did not formally assess whether the lists of key elements for their summaries were consistent across all five LLMs.

### Future Directions and Conclusions

In this study, submitting PICO questions to publicly available generative AI chatbots using a standardized prompt revealed that chatbots both added and missed key information relative to information scientists’ answers. These findings provide insight into LLMs’ capabilities and their potential utility to complement information scientists’ evidence synthesis processes. We recognize that continued and ongoing evaluation will be necessary to keep pace with the rapid evolution of the field and emergence of new models. A critical next phase of this investigation will be to examine all the additional information provided by the chatbots to determine whether it can be validated and supported by the literature, as this will be especially essential for fully understanding the implications for the LLMs’ use by the public. When clinicians review an answer with unfamiliar or unexpected information, they can ask the information scientist and now chatbots for additional clarifying data and, in most instances, they are able to discern the added value of the information they receive. Still, it is important to note that chatbot users may be subject to what is commonly known as automation bias, the propensity to trust information from automated systems without further investigation [42]. Thus, the need to assess the validity of the entire content provided by the answers of the LLM tools as well as the information scientists. Additionally, further investigation into how the public understands and interacts with LLMs to find health information may be needed, as studies have found that LLMs’ performance in answering health-related queries was rated higher when prompted directly by researchers than when prompted by members of the public [44,45].

Although not formally analyzed in this study, we observed that four of the LLMs (all except DeepSeek) commonly included direct links to supporting articles and websites in their answers. In contrast, the answers we previously analyzed from VUMC’s aiChat tool only included in-text references with no direct links, many of which could not be verified to exist [18]. Being able with new versions of the LLMs to directly access and consult cited references is an improvement in facilitating users’ ability to verify chatbots’ answers and seek additional context. A future planned analysis will conduct a more in-depth examination of the references cited by all five LLMs.

## Acknowledgements

This research was developed through the training and support provided by the Medical Library Association’s Research Training Institute (RTI). The authors would like to acknowledge Spencer DesAutels for his assistance with data visualization.

## Data Availability Statement

All data produced in the present study are available upon reasonable request to the authors.

## Author Contributions Statement

Mallory N. Blasingame: Methodology; investigation; data curation; formal analysis; visualization; writing–original draft; writing–review and editing. Taneya Y. Koonce: Methodology; investigation; data curation; formal analysis; writing–original draft; writing– review and editing. Annette M. Williams: Methodology; investigation; data curation; writing– review and editing. Jing Su: Methodology; investigation; visualization; writing–review and editing. Dario A. Giuse: Methodology; investigation; writing original draft; writing–review and editing. Poppy A. Krump: Methodology; investigation; writing–review and editing. Nunzia B. Giuse: Conceptualization; methodology; investigation; formal analysis; visualization; writing–original draft; writing–review and editing; supervision.

