## Appendix B for "Comparing Five Generative AI Chatbots’ Answers to LLM-Generated Clinical Questions with Medical Information Scientists’ Evidence Summaries"

**Appendix B. List of PICO Questions**

| Question number | Question | LLM question source | Question category |
|---|---|---|---|
| 1 | In patients with type 2 diabetes and moderate renal impairment, does the use of SGLT2 inhibitors compared to DPP-4 inhibitors lead to a greater reduction in cardiovascular events? | ChatGPT | Treatment |
| 2 | Among pregnant women in their first trimester, how does maternal obesity, compared to normal maternal weight, affect the risk of developing gestational diabetes? | ChatGPT | Etiology |
| 3 | In patients diagnosed with early-stage non-small cell lung cancer, how does the presence of lymphovascular invasion on pathology reports compare to its absence in predicting 5-year survival rates? | ChatGPT | Prognosis |
| 4 | In patients with acute myocardial infarction, does a percutaneous coronary intervention (PCI) strategy compared to a conservative medical management strategy improve long-term survival? | Google Gemini | Treatment |
| 5 | In adult patients with recurrent urinary tract infections (UTIs), does a history of previous antibiotic use increase the risk of developing antibiotic-resistant UTIs? | Google Gemini | Etiology |
| 6 | What is the long-term prognosis of a patient with newly diagnosed idiopathic pulmonary fibrosis (IPF) and a forced vital capacity (FVC) of 50% predicted? | Google Gemini | Prognosis |
| 7 | In patients with chronic lower back pain, how effective is physical therapy compared to corticosteroid injections in reducing pain and improving mobility? | Microsoft Copilot | Treatment |
| 8 | For patients with type 1 diabetes, how does early viral infection exposure compare to no early viral infection in contributing to the onset of the disease? | Microsoft Copilot | Etiology |
| 9 | In patients with early-stage non-small cell lung cancer, how does the presence of circulating tumor cells affect overall survival compared to those without detectable circulating tumor cells? | Microsoft Copilot | Prognosis |
| 10 | For patients with chronic lower back pain, is physical therapy more effective than non-steroidal anti-inflammatory drugs (NSAIDs) in improving pain and functional mobility over six months? | ChatGPT | Treatment |

| | | | |
|---|---|---|---|
| 11 | In children under the age of five, does early exposure to secondhand smoke, compared to no exposure, increase the likelihood of developing asthma or other chronic respiratory conditions? | ChatGPT | Etiology |
| 12 | In patients with newly diagnosed multiple myeloma, how does the detection of high-risk cytogenetic abnormalities compared to standard-risk cytogenetics affect overall survival over a 3-year period? | ChatGPT | Prognosis |
| 13 | For adults with type 2 diabetes mellitus, does the addition of a GLP-1 receptor agonist to standard metformin therapy reduce the risk of major adverse cardiovascular events compared to metformin alone? | Google Gemini | Treatment |
| 14 | In children with autism spectrum disorder (ASD), does exposure to environmental toxins during pregnancy or early childhood increase the risk of developing ASD? | Google Gemini | Etiology |
| 15 | How does the presence of atrial fibrillation affect the prognosis of patients with acute myocardial infarction? | Google Gemini | Prognosis |
| 16 | For adult patients with Type 2 diabetes, does the addition of a GLP-1 receptor agonist to standard metformin therapy improve glycemic control better than adding a DPP-4 inhibitor? | Microsoft Copilot | Treatment |
| 17 | Among individuals diagnosed with celiac disease, what role does genetic predisposition play in comparison to environmental factors such as diet and hygiene practices? | Microsoft Copilot | Etiology |
| 18 | For individuals diagnosed with chronic kidney disease, what is the impact of elevated serum creatinine levels on the progression to end-stage renal disease compared to those with lower serum creatinine levels? | Microsoft Copilot | Prognosis |
| 19 | In patients with newly diagnosed atrial fibrillation, does the initiation of direct oral anticoagulants (DOACs) compared to warfarin reduce the risk of stroke and systemic embolism without increasing major bleeding incidents? | ChatGPT | Treatment |
| 20 | For adults with a history of hypertension, how does high dietary sodium intake, compared to a low-sodium diet, influence the incidence of stroke and cardiovascular events? | ChatGPT | Etiology |

| | | | |
|---|---|---|---|
| 21 | In individuals with chronic hepatitis B infection, how does a persistently elevated ALT level compared to a consistently normal ALT predict the progression to cirrhosis within 5 years? | ChatGPT | Prognosis |
| 22 | In patients with metastatic non-small cell lung cancer (NSCLC) and EGFR mutations, does treatment with a tyrosine kinase inhibitor (TKI) compared to chemotherapy improve overall survival and quality of life? | Google Gemini | Treatment |
| 23 | In patients with chronic obstructive pulmonary disease (COPD), does exposure to air pollution increase the risk of exacerbations and mortality? | Google Gemini | Etiology |
| 24 | What is the risk of cardiovascular events in patients with type 2 diabetes mellitus and a persistent albuminuria level of 30 mg/g? | Google Gemini | Prognosis |
| 25 | In patients with acute ischemic stroke, does mechanical thrombectomy within 6 hours of symptom onset lead to better functional outcomes compared to intravenous thrombolysis alone? | Microsoft Copilot | Treatment |
| 26 | In elderly patients with Alzheimer's disease, how does chronic exposure to air pollution compare with genetic factors in influencing the development of the disease? | Microsoft Copilot | Etiology |
| 27 | In patients with heart failure with reduced ejection fraction, how does the presence of a baseline elevated BNP level affect the risk of hospitalization for heart failure compared to those with lower BNP levels? | Microsoft Copilot | Prognosis |
| 28 | For hospitalized patients with severe community-acquired pneumonia, does the addition of corticosteroids to standard antibiotic therapy reduce mortality and the length of hospital stay compared to antibiotics alone? | ChatGPT | Treatment |
| 29 | In middle-aged adults, does long-term high alcohol consumption, compared to moderate or no alcohol use, increase the risk of developing liver cirrhosis? | ChatGPT | Etiology |
| 30 | For patients who have suffered an initial ischemic stroke, how does the presence of intracranial artery stenosis on imaging compare to no detected stenosis in forecasting the risk of recurrent stroke within 3 years? | ChatGPT | Prognosis |
| 31 | In children with asthma, does the addition of inhaled corticosteroids to inhaled beta-agonists reduce the frequency of acute exacerbations and improve lung function compared to inhaled beta-agonists alone? | Google Gemini | Treatment |

| | | | |
|---|---|---|---|
| 32 | In postmenopausal women, does hormone replacement therapy (HRT) increase the risk of developing breast cancer? | Google Gemini | Etiology |
| 33 | What is the likelihood of disease progression in patients with multiple sclerosis who experience a second clinical relapse within the first two years of diagnosis? | Google Gemini | Prognosis |
| 34 | For postmenopausal women with osteopenia, does daily supplementation with vitamin D and calcium reduce the risk of fractures more effectively than exercise alone? | Microsoft Copilot | Treatment |
| 35 | For children diagnosed with ADHD, what is the link between maternal smoking during pregnancy and the occurrence of ADHD compared to children whose mothers did not smoke during pregnancy? | Microsoft Copilot | Etiology |
| 36 | Among patients with newly diagnosed rheumatoid arthritis, how do anti-cyclic citrullinated peptide (anti-CCP) antibody levels predict the likelihood of joint damage progression compared to patients without elevated anti-CCP antibody levels? | Microsoft Copilot | Prognosis |
| 37 | In elderly patients with symptomatic knee osteoarthritis, does hyaluronic acid injection improve pain and joint function more effectively than standard oral acetaminophen? | ChatGPT | Treatment |
| 38 | In adolescents, does frequent consumption of high-sugar beverages, compared to minimal or no intake, increase the risk of developing non-alcoholic fatty liver disease? | ChatGPT | Etiology |
| 39 | In patients with early-stage Alzheimer's disease, how does the presence of significant hippocampal atrophy on MRI compared to minimal atrophy influence the rate of cognitive decline over 5 years? | ChatGPT | Disease Prognosis |
| 40 | In patients with chronic obstructive pulmonary disease (COPD), does the addition of long-acting muscarinic antagonist (LAMA) to inhaled corticosteroids improve lung function, quality of life, and reduce exacerbations compared to inhaled corticosteroids alone? | Google Gemini | Treatment |
| 41 | In adults with obesity, does a diet high in processed foods increase the risk of developing type 2 diabetes mellitus? | Google Gemini | Etiology |

| | | | |
|---|---|---|---|
| 42 | How does the presence of a family history of Alzheimer's disease impact the risk of developing dementia in individuals with mild cognitive impairment? | Google Gemini | Prognosis |
| 43 | In children with acute otitis media, does the use of antibiotic therapy shorten the duration of symptoms compared to watchful waiting? | Microsoft Copilot | Treatment |
| 44 | In adolescents with major depressive disorder, how does experiencing early childhood trauma compare to genetic predisposition in influencing the development of the condition? | Microsoft Copilot | Etiology |
| 45 | In patients with acute ischemic stroke, how does the presence of early ischemic changes on initial CT scans predict long-term functional outcomes compared to those without such changes? | Microsoft Copilot | Prognosis |
