## Appendix C for "Comparing Five Generative AI Chatbots’ Answers to LLM-Generated Clinical Questions with Medical Information Scientists’ Evidence Summaries"

**Appendix C. Prompt 1: PICO Question Generation**

*Prompt 1a. Treatment Questions*

#CONTEXT#
I am a medical librarian at a major academic health sciences center. In my team, our members provide evidence-based filtered summaries of the biomedical literature for use in patient care.

#OBJECTIVE#
Your task is to generate a list of five clinical questions that a physician might have while taking care of patients. The questions should be in PICO (Patient, Intervention, Comparison, Outcome) format. These questions should address the category of Treatment.

#STYLE#
The questions should be written in a style likely to be used by a practicing physician.

#TONE#
Maintain a professional and direct tone.

#AUDIENCE#
The audience of the questions is a medical librarian who will answer each inquiry with a search of the literature and filtered evidence summary. Assume the clinical librarian is experienced and familiar with the medical topic at hand.

#RESPONSE FORMAT#
Do not individually separate the PICO elements but rather provide a question that seamlessly includes its elements. Additionally, clearly differentiate each of the five questions you provide.

*Prompt 1b. Etiology Questions*

#CONTEXT#
I am a medical librarian at a major academic health sciences center. In my team, our members provide evidence-based filtered summaries of the biomedical literature for use in patient care.

#OBJECTIVE#
Your task is to generate a list of five clinical questions that a physician might have while taking care of patients. The questions should be in PICO (Patient, Intervention, Comparison, Outcome) format. These questions should address the category of Disease Etiology, defined as "the cause or causes of a disease."

#STYLE#
The questions should be written in a style likely to be used by a practicing physician.

#TONE#
Maintain a professional and direct tone.

#AUDIENCE#
The audience of the questions is a medical librarian who will answer each inquiry with a search of the literature and filtered evidence summary. Assume the clinical librarian is experienced and familiar with the medical topic at hand.

#RESPONSE FORMAT#
Do not individually separate the PICO elements but rather provide a question that seamlessly includes its elements. Additionally, clearly differentiate each of the five questions you provide.

*Note*: The definition for etiology was taken from MedlinePlus [1].

*Prompt 1c. Prognosis Questions*

#CONTEXT#
I am a medical librarian at a major academic health sciences center. In my team, our members provide evidence-based filtered summaries of the biomedical literature for use in patient care.

#OBJECTIVE#
Your task is to generate a list of five clinical questions that a physician might have while taking care of patients. The questions should be in PICO (Patient, Intervention, Comparison, Outcome) format. These questions should address the category of Disease Prognosis, defined as "likely outcome or course of a disease; the chance of recovery or recurrence." In the context of Disease Prognosis, the intervention does not have to be a treatment but could be a lab result, EKG/imaging, or other indicator.

#STYLE#
The questions should be written in a style likely to be used by a practicing physician.

#TONE#
Maintain a professional and direct tone.

#AUDIENCE#
The audience of the questions is a medical librarian who will answer each inquiry with a search of the literature and filtered evidence summary. Assume the clinical librarian is experienced and familiar with the medical topic at hand.

#RESPONSE FORMAT#
Do not individually separate the PICO elements but rather provide a question that seamlessly includes its elements. Additionally, clearly differentiate each of the five questions you provide.

*Note*: The definition for prognosis was taken from the National Cancer Institute [2].

*Follow-up prompt for cases where a question was excluded (e.g., duplicate questions):*

Can you generate one more question in this category?

*Note:* All prompts were adapted from a prompt used in our team's previous study [3], which was developed based on the COSTAR framework [4].
