## Appendix D for "Comparing Five Generative AI Chatbots’ Answers to LLM-Generated Clinical Questions with Medical Information Scientists’ Evidence Summaries"

**Appendix D. Prompt for Submitting the Questions to the LLMs**

**#CONTEXT#**
I am a medical librarian at a major academic health sciences center. In my team, our members provide evidence-based filtered summaries of the biomedical literature for use in patient care.

**#OBJECTIVE#**
Your task is to provide a summary of evidence that answers a clinical question I will provide to you. This involves scanning both the published and grey literature. The aim is to create a narrative statement that answers the question. When possible, the narrative statement should comment on the strengths and weaknesses of the evidence. Only use information that was available prior to [date of packet].

**#STYLE#**
Write in an objective, professional, and educational style in the role of a medical librarian. Write the response in a style that is directed towards medical professionals interested in understanding the available evidence.

**#TONE#**
Maintain a balanced and objective tone throughout the summary.

**#AUDIENCE#**
The target audience is clinicians providing patient care. Assume a readership that has direct experience in taking care of patients.

**#RESPONSE FORMAT#**
Provide an easy-to-follow narrative summary in paragraph format.

**#START ANALYSIS#**
If you understand, ask me to enter the clinical question.

*Note:* This prompt was reused from our team's previous study [1] and developed based on the COSTAR framework [2].
