## Appendix E for "Comparing Five Generative AI Chatbots’ Answers to LLM-Generated Clinical Questions with Medical Information Scientists’ Evidence Summaries"

**Appendix E. Prompt for Obtaining the Key Elements**

#CONTEXT#
Information scientist with medical librarianship training answering a clinical question.

#OBJECTIVE#
I will provide a clinical question and its answer. Your task is to extract only the key elements from the answer.

#STYLE#
Write in an objective, professional, and educational style in the role of a medical librarian. Write the response in a style that is directed towards medical professionals interested in understanding the available evidence.

#TONE#
Maintain a balanced and objective tone.

#AUDIENCE#
The target audience is clinicians providing patient care. Assume a readership that has direct experience in taking care of patients.

#RESPONSE FORMAT#
A numbered list can be used when more than one key element is identified.

#START ANALYSIS#
If you understand, ask me to enter the clinical question and narrative response.

*In cases where the submitted narrative response was longer than the chatbot's word limit, the following text was added to the end of the prompt:*

There will be two parts to the response. Please do not provide the elements until both parts are provided.

*Note:* All prompts were adapted from a prompt used in our team's previous study [1], which was developed based on the COSTAR framework [2].
